# Multimodal Ultrasound and Clinical Data for Pediatric Appendicitis: Evaluating Label-Construction Circularity in the Regensburg Benchmark

**DOI:** 10.64898/2026.09.30.26364432

**Authors:** Md. Mahbubul Islam, Ainur Yerkos

## Abstract

On the public Regensburg Pediatric Appendicitis Dataset, several recent studies report near-ceiling diagnostic performance using radiologist-interpreted ultrasound findings as tabular model inputs, without processing the underlying images. This raises a question worth answering carefully: how much of that performance reflects genuine diagnostic information, and how much reflects the way this dataset’s outcome labels were constructed? We defined four modeling arms - an image-only arm (a backbone-matched BiomedCLIP pipeline as the primary comparator, with an originally-planned ResNet18 model retained as a secondary baseline), a clinical/laboratory-only model (XGBoost), a radiologist-findings model (Random Forest, treated as a reference built from expert-recorded features rather than a live expert comparator), and a multimodal model fusing frozen BiomedCLIP image embeddings with clinical/laboratory data - and compared them using paired DeLong tests, bootstrap confidence intervals, TOST equivalence testing, and pooled repeated cross-validation. The multimodal model (pooled AUC 0.894) showed significantly higher discrimination than the backbone-matched image-only (0.789, *p <* 0.0001) and clinical-only (0.855, *p <* 0.0001) models. Against the radiologist-findings model (0.934), it initially fell short (*p* = 0.034). We then found that the dataset’s own rule for labeling non-surgically-managed patients uses appendix diameter - one of the radiologist-findings predictors - and, separately, the Alvarado and Pediatric Appendicitis Scores - both retained as clinical/multimodal predictors. Removing appendix diameter dropped the radiologist-findings AUC to 0.790; removing the two scores from the multimodal model changed its AUC only marginally, from 0.894 to 0.890, a difference not statistically distinguishable from no change (*p* = 0.25). Comparing these two corrected models directly, the multimodal model remained significantly higher (0.890 vs. 0.790, *p* = 6.2 × 10^−6^). A supplementary check found that 137 of 578 patients (24%) have ultrasound images containing burned-in caliper measurements; on a matched comparison holding the patient set fixed, excluding these images significantly reduced both the image-only and multimodal models’ AUC, indicating a second, image-level information pathway that our design cannot fully attribute to on-screen numeric values versus other differences between marked and unmarked views. An exploratory check that combined both corrections - restricting the doubly-corrected multimodal model to annotation-free images on the smaller, matched patient subset – found a smaller, non-significant difference against the diameter-removed radiologist-findings model (*p* = 0.176), so our main doubly-corrected comparison should not be read as controlling for this second pathway. The multimodal model showed higher discrimination than the unimodal automated baselines on this benchmark, significantly so in pooled repeated cross-validation; its apparent shortfall against radiologist-derived features is highly sensitive to the dataset’s outcome-definition procedure, although the exact fraction of that shortfall attributable to label-construction dependence, as opposed to genuine diagnostic information, cannot be determined from these data alone.

## 1 Introduction

A child arrives with right-lower-quadrant pain, and someone has to decide, usually within a few hours, whether this is appendicitis. The Alvarado Score [1] and the Pediatric Appendicitis Score [2] give a clinical read; white cell count and CRP add a laboratory one; and in most centers an ultrasound contributes a third - a radiologist looks for a non-compressible, enlarged appendix, a target sign, an appendicolith, free fluid, and factors this into the diagnosis. That step depends on a radiologist experienced with pediatric ultrasound being available, which is not guaranteed outside specialized centers, and this has motivated machine-learning work aimed at automating part of the pathway.

The public Regensburg Pediatric Appendicitis Dataset [3, 4] - clinical, laboratory, and ultrasound-image data from children admitted with suspected appendicitis in Regensburg, Germany - has become a widely used public benchmark for this problem. Marcinkevičs et al. [5], who built and released the dataset, trained concept bottleneck models that predict the same sonographic findings a radiologist would (target sign, free fluid, wall-layer status) directly from the ultrasound images, then predict diagnosis from those *predicted* findings - a fully automated pipeline, reaching an AUROC of 0.80. Using the radiologist’s own recorded findings instead of predicting them from pixels typically yields a substantially higher number, which is an expected cost of automation rather than a surprising one.

Two more recent studies on this dataset report considerably higher performance, and the reason is worth stating precisely. Kendall et al. [6] benchmarked a range of classifiers and feature-selection methods with and without what they term “ultrasound image-descriptive features,” reporting an AUROC of 0.993 for diagnosis with those features included, against 0.87-0.88 without them. Navaei et al. [7] report 99.36% accuracy for a gradient-boosting model whose only surviving imaging-derived predictor after feature selection is appendix diameter. Neither study processes ultrasound pixel data with a convolutional network or other automated image-feature extractor - Kendall et al. describe this explicitly as future work - so what each terms an “imaging” contribution is a small number of structured fields a radiologist recorded while reading the scan: the same fields Marcinkevičs et al. had to predict *from* the pixels to build a fully automated pipeline. This raises a specific question: are these findings simply strong predictors, or does the way this dataset’s labels were constructed make some of them partly circular with the outcome they predict?

We find evidence for the latter, and it turns out to be broader than a single feature. The dataset’s rule for labeling non-surgically-managed patients (Section 2.2) assigns the diagnosis label using appendix diameter together with the Alvarado Score and the Pediatric Appendicitis Score. Appendix diameter is a predictor in the radiologist-findings model; the two scores are retained, as legitimate clinical variables, in the clinical-only and multimodal models. Using any of these to predict a label that was partly built from them is not purely correlational. We are not aware of a prior study on this benchmark that quantifies this or asks what correcting for it does to comparisons between automated and radiologist-derived models.

This benchmark-specific problem is an instance of a broader, increasingly well-documented category of failure in medical machine learning. Systematic reviews of clinical prediction models have repeatedly identified data leakage between predictors and outcome labels as a recurring, often undisclosed source of inflated performance estimates [8], and a growing body of work on shortcut learning shows that models can achieve high apparent accuracy by exploiting incidental, non-pathological signal - acquisition artifacts, embedded annotations, or site-specific conventions - rather than the clinical feature a study intends to measure [9, 10]. The label-construction dependence we describe here is a shortcut of the first kind, arising from how the reference standard itself was built; the burned-in caliper measurements we examine separately (Section 3.7) are a shortcut of the second kind, arising from incidental information embedded directly in the imaging data. To our knowledge, this is the first work to document either pathway, let alone both together, on the Regensburg Pediatric Appendicitis Dataset.

This paper makes four contributions. First, we use a medical-domain-pretrained vision-language encoder (BiomedCLIP [11]), chosen over a generic self-supervised encoder (DINOv2 [12]) using the same patient-level pooled cross-validation procedure, as the automated image backbone, and we evaluate it with a matched image-only baseline rather than pairing it against a differently-built comparator. Second, we keep automated inputs (raw images, clinical/laboratory data) and radiologist-interpreted findings separated across four modeling arms, verified in code. Third, we evaluate everything with a model reporting and evaluation framework informed by TRIPOD+AI [13] - paired DeLong tests [14], bootstrap confidence intervals, TOST equivalence testing, and formal power analysis. Fourth, and most centrally, we identify and quantify label-construction circularity in *both* the radiologist-findings model (via appendix diameter) and the multimodal/clinical models (via the Alvarado and Pediatric Appendicitis Scores), correct for it on both sides, and report what survives.

## 2 Materials and Methods

### 2.1 Dataset and Cohort Construction

We used the Regensburg Pediatric Appendicitis Dataset, obtained from Zenodo (version 1.01, DOI 10.5281/zenodo.7669442) [3]. The raw tabular file holds 782 patient records across 58 variables - demographics, clinical examination findings, laboratory results, the Alvarado and Pediatric Appendicitis Scores, radiologist-interpreted ultrasound findings, and outcome labels (diagnosis, management, severity) - with ultrasound images (1-15 B-mode views per patient, BMP format) provided separately. Table 1 summarizes the two dataset releases used in this study: version 1.01 supplied the tabular clinical/outcome data and the image archive used for all modeling, while version 1.03 was used only for the independent verification, described below, that no larger public image archive exists.

**Table 1.**
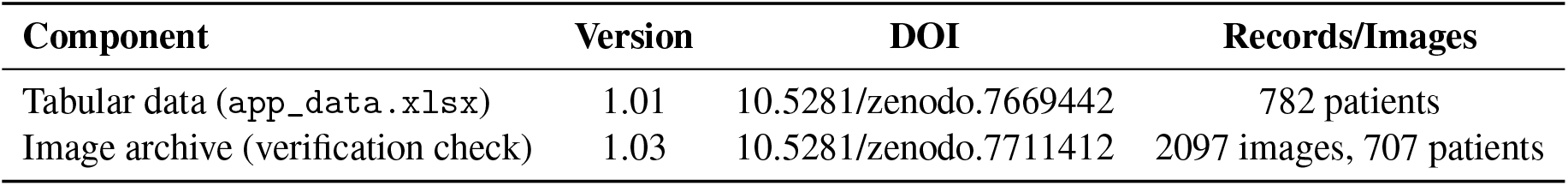
Dataset components and versions used in this study.

| Component | Version | DOI | Records/Images |
| --- | --- | --- | --- |
| Tabular data (app_data.xlsx) | 1.01 | 10.5281/zenodo.7669442 | 782 patients |
| Image archive (verification check) | 1.03 | 10.5281/zenodo.7711412 | 2097 images, 707 patients |

We checked, before any modeling, whether a larger public image set exists that we might simply be missing: version 1.03’s image archive was downloaded and compared against ours patient-by-patient, and the two are identical (2097 images, 707 identifiers in both). No larger public image archive was identified within the dataset releases we evaluated.

Two patients had a missing or unmapped diagnosis label and were dropped, leaving 780 with a valid outcome; 21 more lacked a valid patient identifier, leaving 759 with both. Of these, 578 had at least one matching ultrasound image in the public release; the remaining 181 did not, and 174 of them had an ultrasound performed clinically per the US_Performed field, indicating the scans exist but were not included in the public release. The primary analysis (Sections 3.2-3.3) uses this 578-patient image-linked cohort, split at the patient level into 462 training and 116 test patients from a single fixed split file used across every model. The full 759-patient cohort is used separately in Section 3.9 to check the two image-independent arms on the larger group. Figure 1 shows this funnel.

**Figure 1.**
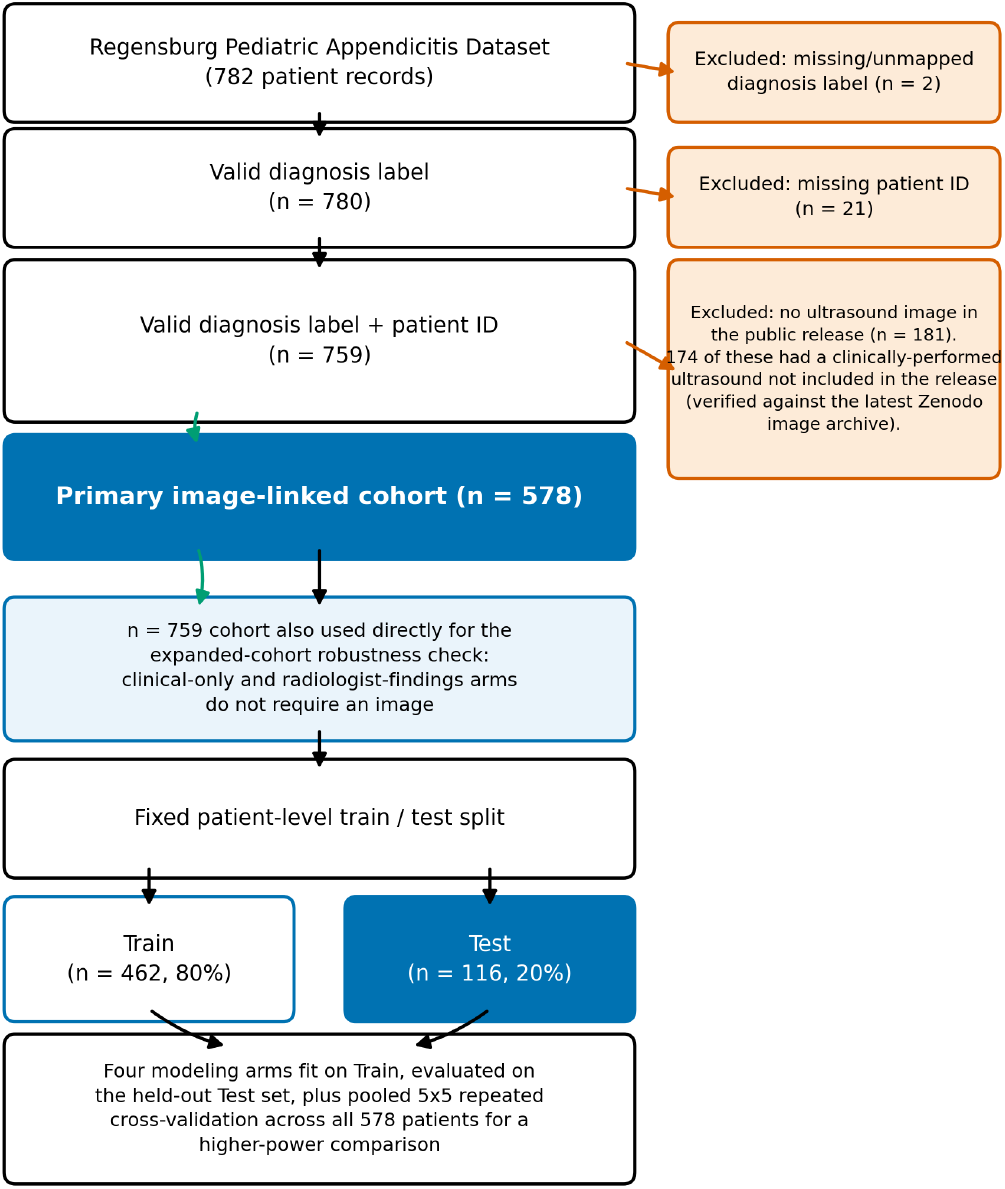
Patient inclusion flow, from the raw 782-record dataset to the primary 578-patient image-linked cohort and its 462/116 train/test split, and to the 759-patient cohort used for the expanded-cohort robustness check.

### 2.2 Ground-Truth Labeling and Label-Construction Circularity

Surgically-treated patients have a histologically confirmed diagnosis. Patients managed conservatively do not, and Marcinkevičs et al. [5] (their Section 2.1) assigned them a label by rule: a conservatively-managed patient is labeled appendicitis-positive if the appendix diameter on ultrasound was at least 6 mm *and* either the Alvarado Score or the Pediatric Appendicitis Score was at least 4 - described by the original authors as “only a proxy for the ground-truth disease status.” Formally, writing *D*_*i*_, *AS*_*i*_, and *PAS*_*i*_ for patient *i*’s appendix diameter, Alvarado Score, and Pediatric Appendicitis Score, and restricting to the conservatively-managed subgroup:

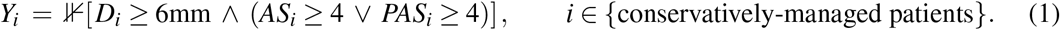

Equation (1) makes the circularity concrete: *D*_*i*_, *AS*_*i*_, and *PAS*_*i*_ are simultaneously arguments of the label-generating function *Y*_*i*_ and, in Section 2.4 onward, components of the feature vector **x**_*i*_ offered to a model trained to predict *Y*_*i*_ from **x**_*i*_. For surgically-treated patients, by contrast, *Y*_*i*_ is set by histology and is not a function of *D*_*i*_, *AS*_*i*_, or *PAS*_*i*_ at all - the same symbol *Y*_*i*_ therefore denotes an outcome constructed by two different processes depending on management pathway, which is the dependency this study investigates.

This rule touches a large part of the cohort. Of 578 patients, 346 (60%) were managed conservatively, and 152 of those are labeled appendicitis-positive; 149 of the 152 (98%) satisfy the rule exactly as recorded in the released tabular data. We inspected the three apparent exceptions individually: two have appendix diameter above the 6 mm threshold but a missing (not recorded) Alvarado and Pediatric Appendicitis Score, so the rule cannot be evaluated for them rather than being violated; the third has an appendix diameter well above threshold (10.7 mm) but both scores recorded below 4, and does not satisfy the stated rule on the data as released. We cannot determine from the available data whether this reflects an additional labeling criterion not documented in the original publication, a data-entry inconsistency, or a clinical override; we report it as an unexplained discrepancy rather than resolving it. Table 2 lays out which variables are involved in this rule and where each is used downstream, since the dependency is not limited to one feature or one modeling arm: appendix diameter enters the radiologist-findings model, and the Alvarado and Pediatric Appendicitis Scores enter both the clinical-only and multimodal models as ordinary clinical variables.

**Table 2.** Label-independence audit: variables involved in the conservative-patient labeling rule and where each is used as a model input.

| Variable | Used in Labeling Rule? | Used in Radiologist-Findings Model? | Used in Clinical/Multimodal Models? |
| --- | --- | --- | --- |
| Appendix Diameter | Yes | Yes | No (excluded) |
| Alvarado Score | Yes | No | Yes |
| Pediatric Appendicitis Score | Yes | No | Yes |
| Other 19 radiologist findings | No | Yes | No (excluded) |

This is directly relevant to Kendall et al. [6], whose best diagnosis model retains appendix diameter and an appendix-visualization flag, and to Navaei et al. [7], whose final model retains appendix diameter as its only imaging-derived predictor; neither discusses this dependency. We quantify it on both sides - the radiologist-findings model by removing appendix diameter, and the clinical/multimodal models by removing both scores - in Sections 3.4 and 3.5. We refer to this phenomenon as label-construction circularity, or outcome-definition dependence, rather than conventional train/test leakage: no information crosses from the test fold into training here; instead, a predictor is partly a component of the target it is used to predict, for a specific subgroup of patients. Where we describe a model below as **label-rule-independent** (rather than simply “label-independent”), this means only that it excludes the specific structured variables identified as components of the published labeling rule (appendix diameter, Alvarado Score, Pediatric Appendicitis Score); it does not mean the model is free of every possible pathway by which expert-derived information could reach it, since the separate image-annotation pathway examined in Section 3.7 remains present in these models’ image inputs.

We also checked a second, independent pathway by which expert-derived information could reach an automated model: whether the ultrasound images themselves contain on-screen measurement calipers or displayed measurement values. The dataset’s own image-naming convention flags a subset of files with an “_M” (measure/marker) suffix, and inspection of a sample confirmed these do contain a visible caliper mark and an on-screen millimeter value; the dataset does not record who placed these markers (radiologist, sonographer, or other operator), so we describe them neutrally as on-screen measurements rather than attributing them to a specific role. Within the 578-patient cohort, 137 patients (24%) have *every* available image tagged this way; a further, larger share have at least one such image among several. We report a sensitivity analysis excluding these images in Section 3.7.

### 2.3 Modeling Arms

We defined four modeling arms on the 578-patient image-linked cohort, all sharing the identical patient-level train/test split so that paired comparisons are valid:

1. **Image-only**. Our primary image-only comparator is a **backbone-matched** model: the identical BiomedCLIP embedding, pooling, and PCA pipeline used in the multimodal arm (Section 2.4), fed to its own XGBoost classifier with no clinical input, so that any multimodal-vs-image-only gain can be attributed to fusion rather than to a backbone difference. We additionally report, as a secondary historical baseline, a ResNet18 convolutional network pretrained on ImageNet and fully fine-tuned on the training images, predicting diagnosis from a single ultrasound frame with patient-level predictions from mean-pooled per-image probabilities; this was our original image-only design and is retained for continuity with our own earlier analysis, not as the primary comparator.
2. **Clinical-only**. An XGBoost classifier [15] on demographic, clinical-examination, laboratory, and scoring variables. All diagnosis-derived columns (Diagnosis_Presumptive, Severity, Management, Length_of_Stay) and all 20 radiologist-interpreted ultrasound-finding columns were excluded, verified in code by an assertion of zero overlap with the radiologist-findings feature list. We separately report a **label-rule-independent** variant with the Alvarado and Pediatric Appendicitis Scores additionally removed (Section 3.5).
3. **Radiologist-findings**. A Random Forest on the 20 radiologist-interpreted ultrasound findings alone (appendix visualization, appendix diameter, free fluid, wall-layer status, target sign, appendicolith, perfusion, perforation, surrounding tissue reaction, abscess and its location, pathological lymph nodes and their location, bowel wall thickening, conglomerate of bowel loops, ileus, coprostasis, meteorism, enteritis, gynecological findings). We refer to this as a **radiologist-derived-feature reference model**, not a live expert comparator: it requires a radiologist’s prior interpretation to exist as structured data, which an automated model cannot assume, and its performance is not a measurement of human diagnostic accuracy.
4. **Multimodal fusion**. Automated image embeddings combined with the same clinical/laboratory features as the clinical-only arm (Section 2.4); no radiologist-interpreted finding enters this arm. We separately report a label-rule-independent variant (Alvarado/Pediatric Appendicitis Scores removed) in Section 3.5.

### 2.4 Image Backbone and Multimodal Fusion Architecture

For the multimodal and backbone-matched image-only arms we used a frozen BiomedCLIP ViT-B/16 encoder [11], pretrained on roughly 15 million biomedical image-caption pairs - a strong frozen biomedical foundation encoder, though not one pretrained specifically on pediatric ultrasound. BiomedCLIP was selected over a frozen DINOv2 ViT-S/14 encoder [12] using the same patient-level pooled cross-validation procedure applied to both backbones: BiomedCLIP came out ahead (pooled AUC 0.894 vs. 0.883 for the multimodal arm). Both stayed fully frozen throughout; an earlier attempt at partially fine-tuning a comparable encoder made things worse (0.744 vs. 0.756 frozen), consistent with there being too few training images (462-759) to fine-tune a vision transformer without overfitting it.

Images were used as released by the dataset, converted to RGB, resized and center-cropped to each encoder’s native input resolution (224 × 224), and normalized with each encoder’s own published channel statistics; center cropping was used only to satisfy each encoder’s fixed input size, not to select a region of interest, so no manual or automated appendix localization was provided to either model. No augmentation and no masking or inpainting of on-screen text, calipers, or other burned-in graphical elements was applied. This means the models had access to whatever on-screen information the original frames contain, including the caliper markings discussed in Section 2.2 and evaluated in Section 3.7; we did not apply pixel-level removal of these elements in the primary analysis.

Each patient’s 1-15 images are encoded independently into 512-dimensional embeddings, then combined into one patient-level embedding by weighting each image’s contribution by the norm of its own embedding - an **L2-norm-weighted pooling**, not a learned attention mechanism in the neural-network sense, despite some similarity in effect. Formally, for a patient with image embeddings **z**_1_, …, **z**_*n*_ *∈* ℝ^512^ (one per available ultrasound frame), the patient-level embedding **z**_*p*_ is

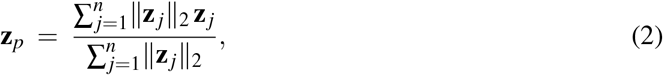

i.e., a convex combination of the per-image embeddings weighted by each embedding’s own Euclidean norm, so that images whose encoder representation is more strongly activated contribute proportionally more to the pooled representation, with no learned parameters governing the weighting. That embedding is reduced to 50 dimensions by PCA, fit on the training set only, and concatenated with the clinical/laboratory feature vector. The result feeds an XGBoost classifier tuned with Optuna (40 trials, Tree-structured Parzen Estimator sampler) via 5-fold stratified group cross-validation on the training set alone. Figure 2 shows the pipeline; the clinical pathway shown there excludes only the diagnosis-derived and radiologist-finding columns by default, with the additional Alvarado/PAS exclusion applied specifically for the label-rule-independent sensitivity variant of Section 3.5.

**Figure 2.**
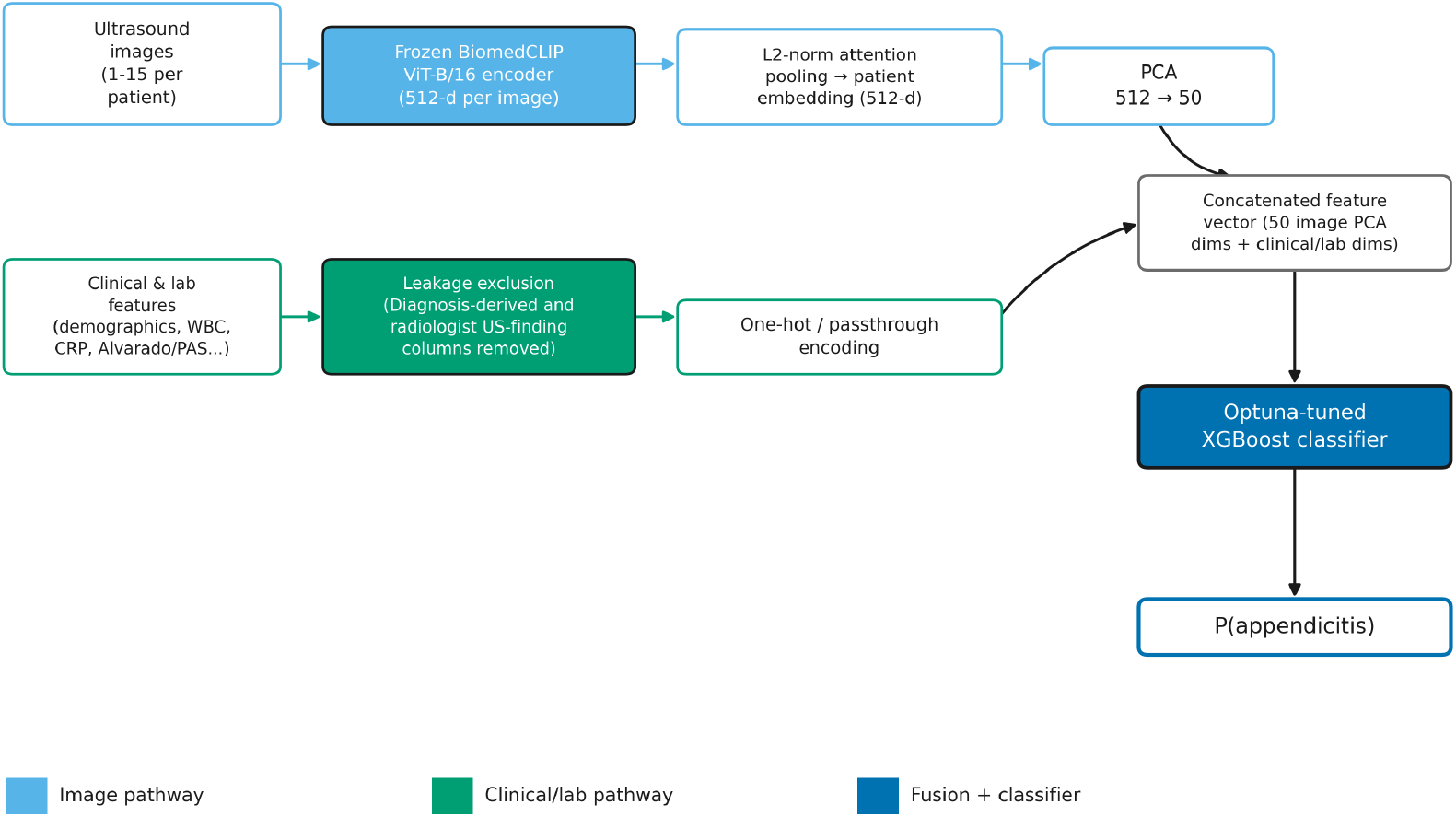
Multimodal fusion architecture: frozen BiomedCLIP image encoder with L2-norm-weighted pooling and PCA, concatenated with feature-filtered clinical/laboratory inputs, classified by an Optuna-tuned XGBoost model.

### 2.5 Statistical Analysis

Throughout this paper we report our own models’ discrimination as AUC (area under the receiver operating characteristic curve); where we cite previously published results, we retain the metric name - AUC or AUROC - exactly as reported by the original authors, since these are the same quantity under different labels. We quantified discrimination with the AUC, with 95% confidence intervals from 1000 patient-level bootstrap resamples, and calibration with the Brier score and calibration curves (10 quantile-based bins per curve); with 116 primary-split patients, these curves are descriptive and not powered to detect small miscalibration. Arms were compared pairwise with DeLong’s test [14] for correlated ROC curves on the same patients. For two models *A* and *B* evaluated on the same *n* patients, with estimated AUCs 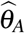 and 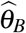 and DeLong-estimated variance-covariance terms 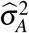, 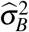, and 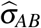, the test statistic is

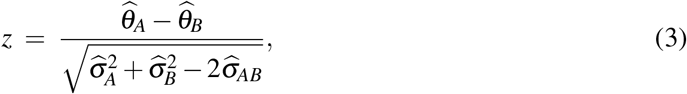

referred to a standard normal distribution to obtain a two-sided *p*-value; the covariance term 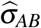 is what makes this test appropriate for paired predictions on the same patients, as opposed to treating the two AUCs as independent. This was supplemented by a Two One-Sided Tests (TOST) procedure at a pre-specified *±*0.05 AUC margin *δ*, which tests the two one-sided hypotheses *H*_01_ : *θ*_*A*_ *−θ*_*B*_ *≤ −δ* and *H*_02_ : *θ*_*A*_ *−θ*_*B*_ *≥ δ* against the alternative that |*θ*_*A*_ *−θ*_*B*_| *< δ*, using the same DeLong standard error as Equation (3); the combined TOST *p*-value is the larger (less significant) of the two one-sided *p*-values, *p*_TOST_ = max(*p*_1_, *p*_2_), and equivalence at the *α* level is concluded only if *p*_TOST_ *< α*. We did not derive the margin *δ* = 0.05 from a clinical-outcome or literature-based justification; we prespecified it as the methodological threshold we considered practically negligible for this benchmark comparison, fixed before any comparison was examined and not fitted to the results - with both one-sided *p*-values reported alongside the combined TOST *p*-value. We also reported the minimum detectable effect size (80% power, two-sided *α* = 0.05, computed from each comparison’s observed DeLong standard error) for every comparison.

Because a single 462/116 split has limited power for the differences of interest, we additionally ran every comparison through pooled repeated cross-validation: 5-fold stratified cross-validation repeated 5 times with different random splits (25 fold-fits per arm), averaging each patient’s out-of-fold probability across whichever repeats held that patient out. Within each of the 25 folds, PCA (and every other fitted preprocessing step: scaling, one-hot encoding) was refit from scratch on that fold’s training patients only and applied unchanged to that fold’s held-out patients, so no fold’s validation patients contributed to their own fold’s PCA basis or feature scaling. Hyperparameters, by contrast, were chosen once by the training-only Optuna search and held fixed across all 25 folds; we did not re-tune hyperparameters within each outer fold (i.e., this is a non-nested design), which trades a small, difficult-to-quantify amount of optimism in the pooled estimates for a substantial reduction in computation, and we flag this explicitly as a limitation (Section 4.3) rather than describing the resulting predictions as fully nested out-of-sample estimates.

Of the comparisons reported, three were pre-specified before any circularity analysis was conducted (image-only vs. multimodal, clinical-only vs. multimodal, multimodal vs. radiologist-findings, each evaluated on both the primary split and pooled cross-validation); the appendix-diameter and Alvarado/PAS sensitivity analyses, the annotation-contamination check, and the expanded-cohort and backbone/ensemble checks were exploratory, motivated by patterns noticed during the primary analysis, and we label them as such throughout.

### 2.6 Sensitivity Analyses

1. **Appendix-diameter circularity**. The radiologist-findings model was refit with appendix diameter removed from its 20-feature input.
2. **Alvarado/PAS circularity**. The clinical-only and multimodal models were refit with the Alvarado and Pediatric Appendicitis Scores removed from their clinical feature set.
3. **Image-annotation contamination**. The backbone-matched image-only and multimodal models were refit using only ultrasound images without a visible caliper/measurement marker, on the subset of 441 patients (of 578) who retained at least one such image after exclusion.
4. **Expanded-cohort robustness**. The clinical-only and radiologist-findings arms were re-evaluated on the full 759-patient cohort.
5. **Backbone and ensemble ablations**. A feature-level BiomedCLIP+DINOv2 ensemble, a prediction-level average of the two backbones, and a three-algorithm ensemble were evaluated against the single-backbone multimodal model (Supplementary Materials).

All analyses used a fixed random seed (42). Code and the exact prediction files behind every number reported here are described in the Data Availability statement.

## 3 Results

### 3.1 Baseline Cohort Characteristics

Table 3 summarizes the 578-patient cohort by diagnosis. Patients with appendicitis were, on average, slightly younger, had more variable BMI, and had markedly higher inflammatory markers, clinical scores, and appendix diameter than those without (all *p <* 0.05). We note that, because appendix diameter and the two clinical scores participate in the labeling rule for conservatively-managed patients (Section 2.2), the sharp separation seen for these three variables in Table 3 is partly definitional rather than solely a biological association to be taken at face value.

**Table 3.** Baseline characteristics of the 578-patient image-linked cohort, by diagnosis.

| Characteristic | Appendicitis (n=382) | No Appendicitis (n=196) | p-Value |
| --- | --- | --- | --- |
| Age, years, mean ± SD | 11.01 ± 3.58 | 11.87 ± 3.32 | 0.009 |
| Sex, male, n (%) | 219 (57.6) | 93 (47.4) | 0.028 |
| BMI, kg/m<sup>2</sup>, mean ± SD | 18.40 ± 4.12 | 19.20 ± 4.48 | 0.026 |
| WBC count, $\times 10^9$/L, mean ± SD | 14.18 ± 5.20 | 10.12 ± 4.57 | < 0.001 |
| CRP, mg/L, mean ± SD | 43.71 ± 68.14 | 10.88 ± 25.96 | < 0.001 |
| Alvarado Score <sup>†</sup>, mean ± SD | 6.67 ± 1.91 | 4.84 ± 2.06 | < 0.001 |
| Pediatric Appendicitis Score <sup>†</sup>, mean ± SD | 5.83 ± 1.86 | 4.45 ± 1.86 | < 0.001 |
| Appendix diameter <sup>†</sup>, mm, mean ± SD | 8.82 ± 2.18 | 5.07 ± 1.17 | < 0.001 |
| Management, surgical, n (%) | 230 (60.2) | 2 (1.0) | - |
| Management, conservative, n (%) | 152 (39.8) | 194 (99.0) | - |

<sup>†</sup> Involved in the diagnostic labeling rule for conservatively-managed patients (Section 2.2).

### 3.2 Per-Arm Performance: Primary Held-Out Split (n=116)

On the primary 116-patient held-out set (Table 4), the radiologist-findings model scored highest (AUC 0.942), followed by the multimodal model (0.856), clinical-only (0.841), the backbone-matched BiomedCLIP image-only model (0.733), and, as a secondary historical baseline, the original ResNet18 image-only model (0.686). We report all downstream primary-split comparisons using the backbone-matched BiomedCLIP image-only model, so that the image-only-vs-multimodal comparison isolates the effect of fusion rather than a backbone difference; the ResNet18 figure is retained in Table 4 for continuity with our earlier analysis only.

**Table 4.** Per-arm performance on the primary held-out test set (n=116).

| Arm | AUC (95% CI) | AP (95% CI) | Brier (95% CI) | Sens.@0.5 | Spec.@0.5 |
| --- | --- | --- | --- | --- | --- |
| Image-only (BiomedCLIP, matched, primary) | 0.733 (0.636-0.826) | 0.833 (0.727-0.915) | 0.196 (0.162-0.234) | 0.818 | 0.487 |
| Image-only (ResNet18, secondary baseline) | 0.686 (0.592-0.789) | 0.814 (0.720-0.892) | 0.245 (0.190-0.298) | 0.727 | 0.538 |
| Clinical-only | 0.841 (0.755-0.917) | 0.887 (0.792-0.960) | 0.154 (0.125-0.186) | 0.922 | 0.538 |
| Radiologist-findings | 0.942 (0.890-0.977) | 0.967 (0.933-0.989) | 0.086 (0.055-0.123) | 0.909 | 0.846 |
| Multimodal (BiomedCLIP) | 0.856 (0.776-0.926) | 0.910 (0.834-0.965) | 0.137 (0.101-0.179) | 0.857 | 0.769 |

Table 5 reports the three pre-specified paired comparisons, all using the backbone-matched BiomedCLIP image-only and multimodal models. The multimodal model showed significantly higher discrimination than image-only (*p* = 0.0112). The comparisons against clinical-only (*p* = 0.740) and radiologist-findings (*p* = 0.061) were not significant; the minimum detectable AUC difference here (80% power) was 0.121-0.137, larger than the point estimates themselves, so these two comparisons are inconclusive at this sample size rather than evidence that no difference exists. A TOST equivalence test for multimodal-vs-radiologist-findings, at the pre-specified *±*0.05 margin, gave a one-sided *p* of 0.0015 for the lower bound and 0.781 for the upper bound (combined TOST *p* = 0.781): equivalence was not established, and this comparison remained inconclusive in both directions at n=116.

**Table 5.** Paired DeLong comparisons, primary held-out split (n=116), using the backbone-matched BiomedCLIP image-only model.

| Comparison | AUC (A) | AUC (B) | Diff (A–B) | 95% CI | p-Value |
| --- | --- | --- | --- | --- | --- |
| Image-only vs. Multimodal | 0.733 | 0.856 | −0.124 | (−0.220, −0.028) | 0.0112 |
| Clinical-only vs. Multimodal | 0.841 | 0.856 | −0.014 | (−0.099, 0.070) | 0.7397 |
| Multimodal vs. Radiologist-findings | 0.856 | 0.942 | −0.085 | (−0.174, 0.004) | 0.0607 |

For reference, pairing the original ResNet18 image-only model against the same 0.856 multi-modal figure gives diff = −0.170, *p* = 0.0049 - the qualitative conclusion (multimodal shows higher discrimination than image-only) is unchanged by the choice of image-only backbone, but only the BiomedCLIP-matched comparison isolates the fusion effect specifically.

### 3.3 Pooled Repeated Cross-Validation (n=578)

Pooling across all 578 patients (Table 6) shrank the minimum detectable difference to 0.02-0.05. Using the backbone-matched BiomedCLIP image-only model (0.789) rather than the ResNet18 model, both previously inconclusive comparisons resolved (Table 7): the multimodal model showed significantly higher discrimination than the backbone-matched image-only model (*p <* 0.0001) - evidence consistent with a multimodal fusion benefit rather than merely a backbone effect, though see Section 4.3 on the non-nested cross-validation design underlying this estimate - and the clinical-only model as well (*p <* 0.0001). The radiologist-findings comparison also resolved, in the radiologist-findings model’s favor (*p* = 0.0337).

**Table 6.** Per-arm performance, pooled repeated cross-validation (n=578).

| Arm | Pooled AUC (95% CI) |
| --- | --- |
| Image-only (BiomedCLIP, matched) | 0.789 (0.752-0.824) |
| Clinical-only | 0.855 (0.823-0.884) |
| Multimodal (BiomedCLIP) | 0.894 (0.868-0.917) |
| Radiologist-findings | 0.934 (0.907-0.956) |

**Table 7.** Paired DeLong comparisons, pooled repeated cross-validation (n=578), using the backbone-matched image-only model.

| Comparison | AUC (A) | AUC (B) | Diff (A–B) | 95% CI | p-Value |
| --- | --- | --- | --- | --- | --- |
| Image-only vs. Multimodal | 0.789 | 0.894 | −0.105 | (−0.141, −0.069) | < 0.0001 |
| Clinical-only vs. Multimodal | 0.855 | 0.894 | −0.039 | (−0.056, −0.022) | < 0.0001 |
| Multimodal vs. Radiologist-findings | 0.894 | 0.934 | −0.039 | (−0.075, −0.003) | 0.0337 |

At this point the reading is: multimodal AI shows significantly higher discrimination than unimodal AI (on a fair, backbone-matched basis), and the radiologist-findings model still scores highest. We next examine how much of that remaining gap survives once known label-construction dependencies are removed from both sides.

### 3.4 The Appendix-Diameter Circularity Effect

Removing appendix diameter from the radiologist-findings model’s 20-feature input dropped its pooled AUC from 0.934 to 0.790 (Table 8) - an observed AUC decrease of 0.144 after removing one predictor. This drop provides a bound on the combined contribution of genuine diagnostic information and label-construction dependence in this feature; it does not by itself establish that the full 0.144 decrease is circularity, since a larger, more inflamed appendix is plausibly informative independent of the labeling rule. Given that 98% of conservatively-managed appendicitis-positive patients satisfy the exact rule involving this measurement, the magnitude and structure of this drop make a non-negligible contribution from label-construction dependence plausible, though we cannot separate that contribution from genuine diagnostic content with the available data. Figure 3 illustrates the mechanism.

**Table 8.**
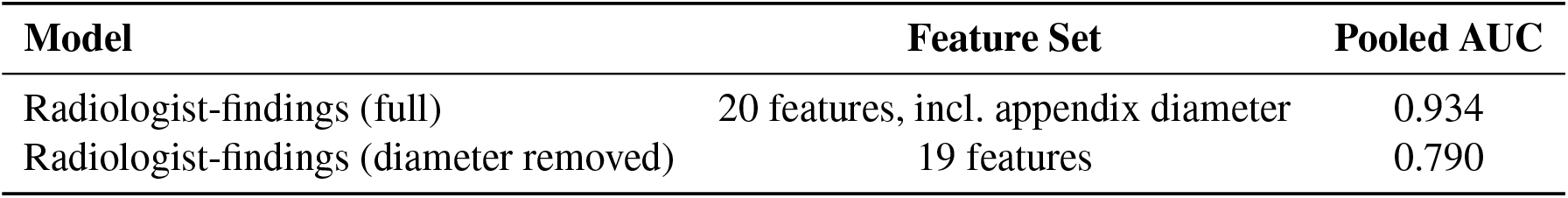
Effect of removing appendix diameter from the radiologist-findings model (pooled, n=578).

| Model | Feature Set | Pooled AUC |
| --- | --- | --- |
| Radiologist-findings (full) | 20 features, incl. appendix diameter | 0.934 |
| Radiologist-findings (diameter removed) | 19 features | 0.790 |

**Figure 3.**
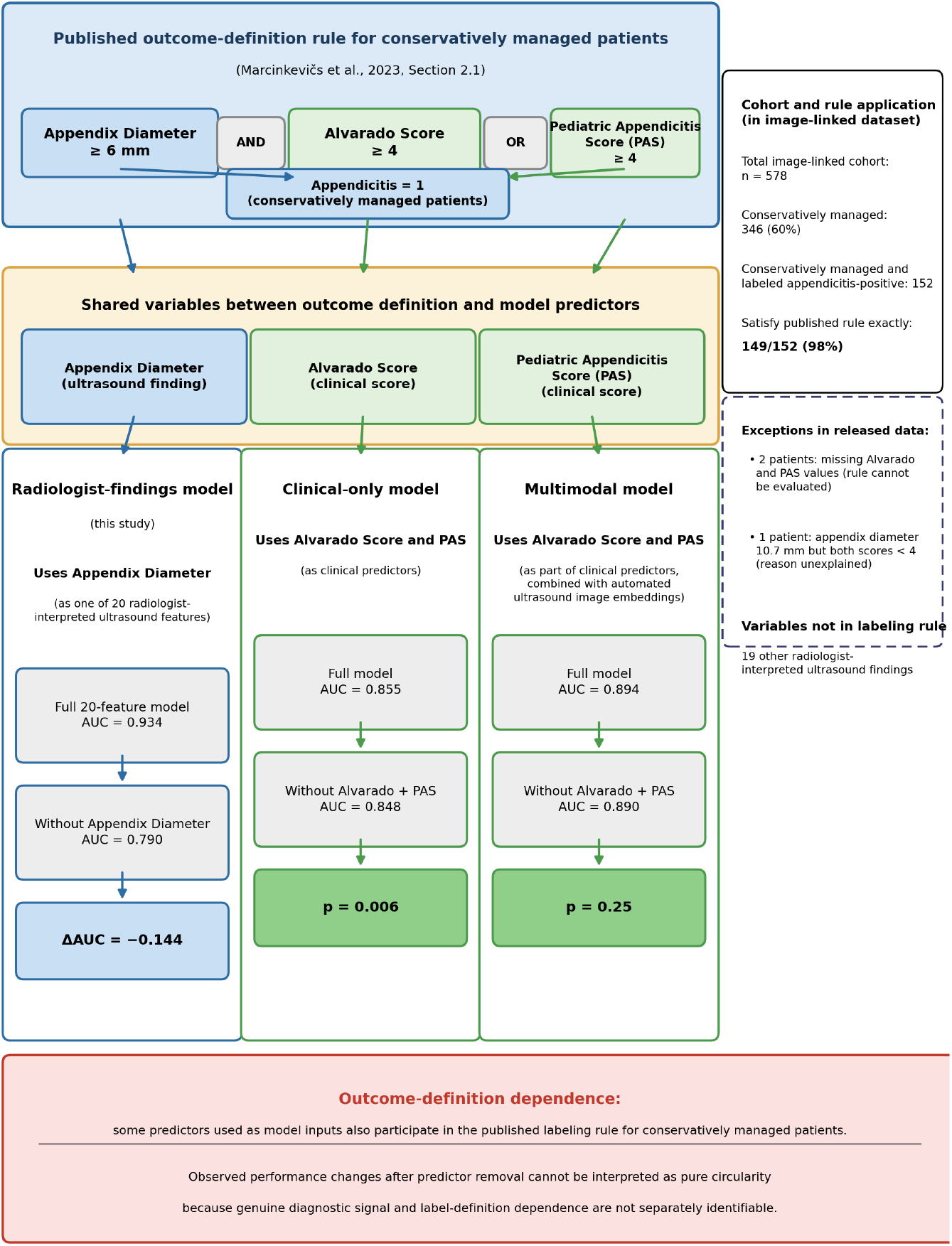
Label-construction circularity and outcome-definition dependence affecting model performance. The dataset’s published diagnostic labeling rule for conservatively-managed patients depends on appendix diameter, the Alvarado Score, and the Pediatric Appendicitis Score (PAS), which are also used as predictors in the corresponding modeling arms. The observed AUC changes after removing these variables quantify sensitivity to these dependencies but cannot distinguish genuine diagnostic information from label-definition dependence.

### 3.5 Alvarado/PAS Circularity in the Clinical and Multimodal Models

Because the labeling rule also involves the Alvarado and Pediatric Appendicitis Scores, we applied the same correction to the clinical-only and multimodal models (Table 9). Removing both scores dropped the clinical-only pooled AUC from 0.855 to 0.848 (*p* = 0.006) - a small but statistically detectable effect - and changed the multimodal pooled AUC from 0.894 to 0.890, which was not statistically significant (*p* = 0.25). A non-significant result at *p* = 0.25 does not itself establish the absence of circularity; it indicates only that this analysis did not detect a statistically significant change. Unlike appendix diameter, these two scores are themselves aggregates of several clinical signs and symptoms rather than a single thresholded measurement, which is one plausible structural reason their removal produced a smaller drop than removing appendix diameter did; we treat this as a plausible contributing explanation rather than a demonstrated one, and we cannot rule out a modest circularity contribution that this comparison lacked power to detect.

**Table 9.** Effect of removing the Alvarado and Pediatric Appendicitis Scores from the clinical-only and multimodal models (pooled, n=578).

| Model | Full AUC | Label-Independent AUC | p-Value |
| --- | --- | --- | --- |
| Clinical-only | 0.855 | 0.848 | 0.006 |
| Multimodal (BiomedCLIP) | 0.894 | 0.890 | 0.25 |

### 3.6 Comparing the Doubly-Corrected Models

For the central comparison of this study, we paired the label-rule-independent multimodal model against the diameter-removed radiologist-findings model - both sides corrected for the specific variables implicated in Section 2.2. The label-rule-independent multimodal model (pooled AUC 0.890) remained significantly higher than the diameter-removed radiologist-findings model (pooled AUC 0.790; diff = +0.100, 95% CI 0.057-0.143, *p* = 6.2 × 10^−6^; Table 10). This is a DeLong superiority result, not a TOST equivalence result: the 95% confidence interval for the difference excludes zero entirely and lies well outside the pre-specified *±*0.05 equivalence margin in the multimodal model’s favor, so a TOST test here (which we do not report, since TOST is designed to test for equivalence, not to confirm a difference) would be uninformative - the DeLong test and its confidence interval already establish the direction and statistical reliability of this result. This doubly-corrected comparison is more conservative than pairing the uncorrected multimodal model against the diameter-removed radiologist-findings model alone (Table 8, diff +0.104, *p* = 2.6 × 10^−6^): both give the same qualitative answer, and the doubly-corrected version does not depend on an asymmetric correction applied to only one arm. We use “doubly-corrected” specifically to mean corrected for the labeling-rule variables identified in Section 2.2 (appendix diameter; Alvarado Score and Pediatric Appendicitis Score); it does not additionally control for the separate image-annotation pathway identified in Section 3.7, which remains present in the multimodal model’s image input here. Section 3.8 reports what happens when both pathways are controlled for at once.

**Table 10.** Doubly-corrected comparison: label-rule-independent multimodal model vs. diameter-removed radiologist-findings model (pooled, n=578).

| Comparison | Diff (A–B) | 95% CI | p-Value |
| --- | --- | --- | --- |
| Multimodal (full) vs. Radiologist-findings (diameter removed) | +0.104 | (0.061, 0.148) | $2.6 \times 10^{-6}$ |
| Multimodal (label-rule-independent) vs. Radiologist-findings (diameter removed) | +0.100 | (0.057, 0.143) | $6.2 \times 10^{-6}$ |

Figure 4 places the primary, pooled, and circularity-corrected comparisons on one scale, including the doubly-corrected comparison from Table 10.

**Figure 4.**
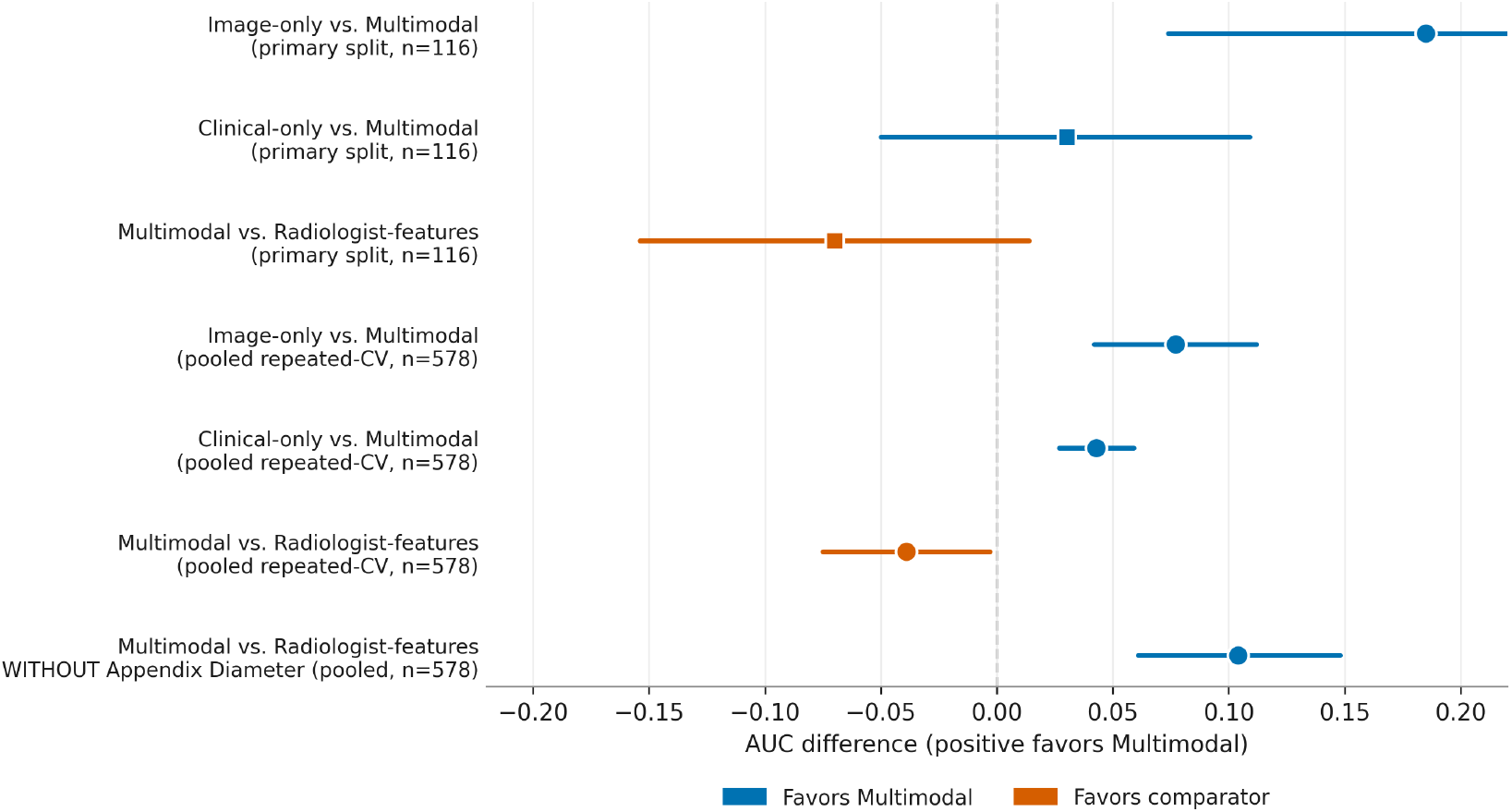
Paired AUC differences across primary and circularity-corrected comparisons (95% CI). AUC differences are expressed as Multimodal − Comparator; positive values favor the multimodal model and negative values favor the comparator.

### 3.7 Image-Annotation Contamination Check

Embedded, non-pathological markings of exactly this kind are a recognized source of shortcut learning in medical imaging [9, 10], and we checked for them directly. Ultrasound images in this dataset are inconsistently marked: 970 of 2097 images (46%) carry a filename suffix indicating an on-screen caliper measurement, and visual inspection confirmed these images display a caliper mark and a millimeter value directly on the frame. Within the 578-patient cohort, 137 patients (24%) have *only* such images, meaning they cannot be evaluated at all once marked images are excluded.

As a first sensitivity check, we rebuilt the backbone-matched image-only and multimodal (label-rule-independent) models using only unmarked images, retaining the 441 patients who had at least one such image; pooled AUC dropped to 0.700 (image-only) and 0.859 (multimodal). This comparison is confounded by both a 24% reduction in cohort size and a possible difference in clinical composition between patients who needed more measurement views and those who did not, so it cannot on its own distinguish an effect of removing caliper information from an effect of evaluating a different, smaller patient set.

We therefore ran a second, better-controlled comparison, restricting *both* the all-images and the annotation-free pipelines’ pooled predictions to the identical 441 patients, so that any difference reflects the image set used rather than a change in cohort (Table 11). On this matched comparison, restricting to annotation-free images significantly reduced AUC for both the image-only model (0.780 to 0.700; diff = 0.080, 95% CI 0.040-0.121, *p* = 8.1 × 10^−5^) and the multimodal model (0.882 to 0.859; diff = 0.023, 95% CI 0.009-0.038, *p* = 0.0018). Because the patient set was held fixed, this analysis reduces the sample-composition confounding present in the first sensitivity analysis and supports a contribution from information associated with differences between marked and unmarked image views. We remain cautious about attributing this specifically to the models reading the displayed numeric value, rather than to marked images more often being closer, better-centered, or otherwise higher-quality views of the appendix independent of any on-screen text; distinguishing these two explanations would require pixel-level masking or inpainting of the caliper region while holding the underlying image constant, which we did not implement. Either explanation means the image-only and multimodal arms’ reported performance may not transfer to a deployment setting where such markings are absent, which we treat as an open limitation (Section 4.3) rather than a resolved question.

**Table 11.** Annotation-clean sensitivity check, paired on the identical 441 patients who retained at least one annotation-free image.

| Arm | All Images | Annotation-Free | Diff (95% CI) | p-Value |
| --- | --- | --- | --- | --- |
| Image-only (BiomedCLIP, matched) | 0.780 | 0.700 | 0.080 (0.040-0.121) | $8.1 \times 10^{-5}$ |
| Multimodal (label-rule-independent) | 0.882 | 0.859 | 0.023 (0.009-0.038) | 0.0018 |

### 3.8 Exploratory Check: Both Correction Pathways Combined

The doubly-corrected comparison in Section 3.6 controls for the labeling-rule variables but not for the image-annotation pathway of Section 3.7; the annotation-clean check controls for the annotation pathway on a reduced, patient-matched subset but was not previously paired against the diameter-removed radiologist-findings model on that same subset. As an exploratory check, we paired the two directly: the label-rule-independent, annotation-free multimodal predictions from Table 11 (pooled AUC 0.859) against the diameter-removed radiologist-findings model’s predictions (Section 3.4), both restricted to the identical 441 patients. The multimodal model remained numerically higher (0.859 vs. 0.822; diff = +0.037, 95% − CI 0.016-0.090), but this difference was not statistically significant (*p* = 0.176), and the confidence interval includes zero.

We report this transparently rather than omit it: the smaller, non-significant estimate here, relative to the doubly-corrected result’s +0.100 (*p* = 6.2 × 10^−6^), is consistent with a combination of reduced statistical power at *n* = 441 (a 24% smaller sample than the primary *n* = 578 pooled analysis) and a possible additional contribution from the annotation pathway to the multimodal model’s advantage; our design cannot distinguish between these two explanations, and this analysis was not powered in advance to detect an effect of this size. We treat this as an open question rather than a resolved one: the central, pre-specified doubly-corrected comparison (Section 3.6) addresses labeling-rule circularity specifically and remains our primary result, but readers should note that a comparison additionally controlling for the image-annotation pathway, though underpowered here, did not reach statistical significance.

### 3.9 Expanded-Cohort Robustness (n=759)

The clinical-only and radiologist-findings arms do not need an image, so both were re-run on the full 759-patient cohort as a robustness check, not an external validation - it remains the same single-center dataset (Table 12). Both held steady (clinical-only: 0.855 vs. 0.857; radiologist-findings: 0.934 vs. 0.929).

**Table 12.** Expanded-cohort robustness check: image-independent arms re-evaluated on the full diagnosed cohort.

| Arm | Pooled AUC (n=578) | Pooled AUC (n=759) |
| --- | --- | --- |
| Clinical-only | 0.855 | 0.857 |
| Radiologist-findings | 0.934 | 0.929 |

### 3.10 Subgroup Performance

We computed the pooled multimodal model’s AUC, with bootstrap 95% confidence intervals, by sex and by age tertile (Table 13). Point estimates were broadly similar across both (0.880-0.912), and the confidence intervals overlap substantially across all five subgroups; we did not formally test for interaction/heterogeneity between subgroups, so this should be read as showing no evidence of a marked subgroup deficit rather than a formal equivalence claim. We did not have reliable imaging-equipment metadata to extend this to a device-level subgroup analysis, which remains a gap.

**Table 13.** Pooled multimodal model performance by sex and age tertile (n=578).

| Subgroup | n | Pooled AUC (95% CI) |
| --- | --- | --- |
| Female | 265 | 0.880 (0.837-0.920) |
| Male | 312 | 0.900 (0.862-0.933) |
| Age tertile 1 (0.0-10.1 y) | 193 | 0.912 (0.872-0.948) |
| Age tertile 2 (10.1-12.8 y) | 192 | 0.888 (0.831-0.934) |
| Age tertile 3 (12.8-17.9 y) | 193 | 0.881 (0.831-0.927) |

### 3.11 Interpretability and Calibration

Figures 5-8 show qualitative Grad-CAM visualizations for the ResNet18 image-only model (four hand-selected cases, not a validated localization metric), impurity-based feature importance for the radiologist-findings model, calibration curves, and decision curve analysis. The multimodal model’s calibration on the primary split sat between the clinical-only and radiologist-findings arms (Brier score 0.137, against 0.154 and 0.086 respectively; Table 4).

**Figure 5.**
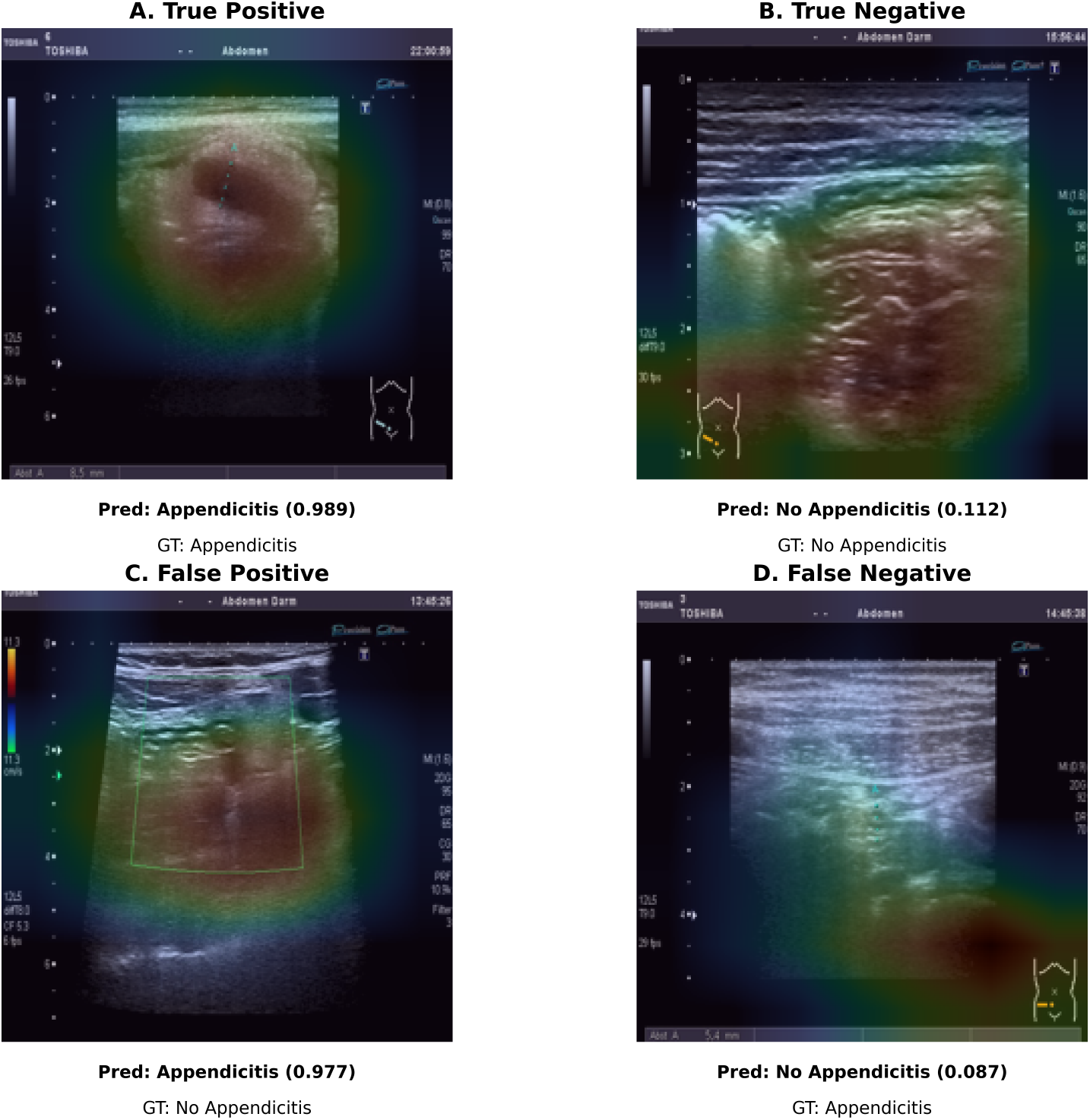
Grad-CAM visualizations of the ResNet18 image-only model on representative true-positive, true-negative, and misclassified cases. These are qualitative examples, not a validated measure of localization accuracy.

**Figure 6.**
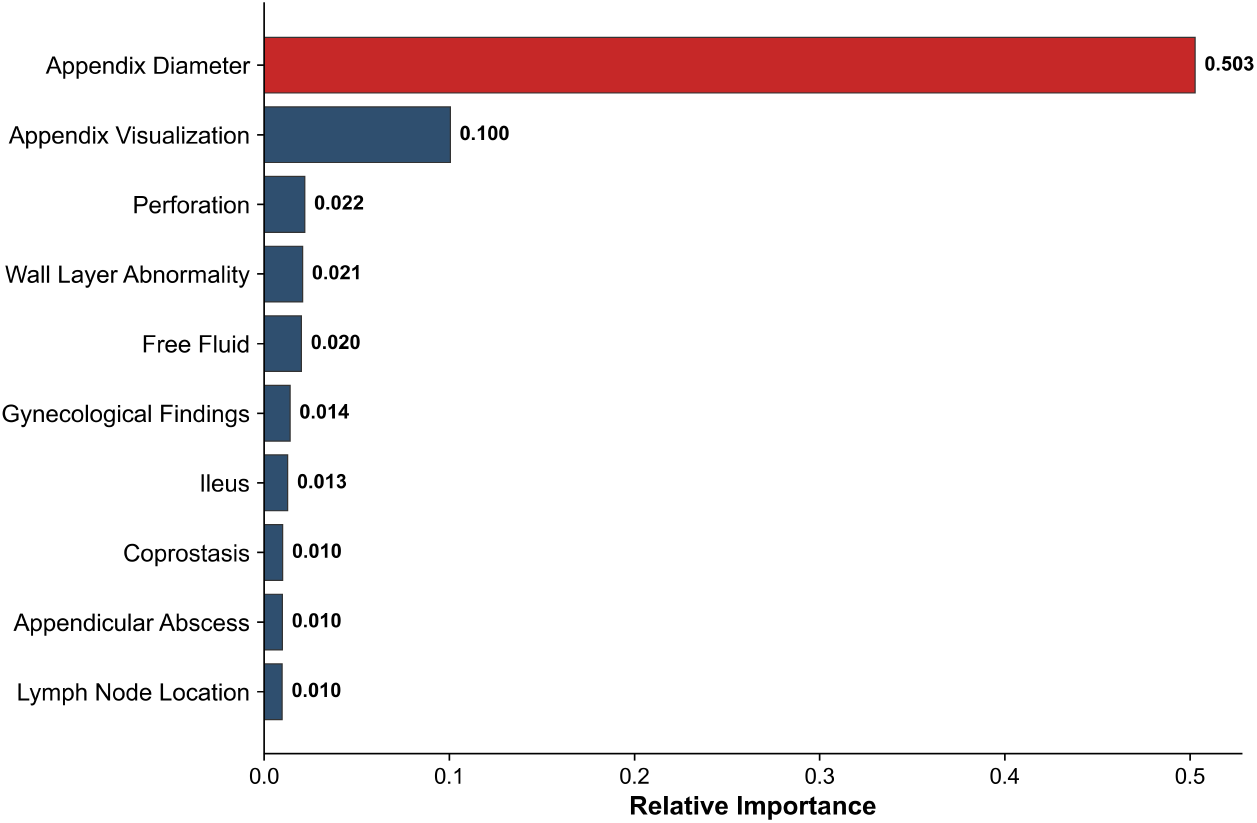
Impurity-based feature importance for the radiologist-findings model. This is a descriptive summary of split contribution within the fitted trees, not a validated measure of each variable’s causal or standalone predictive contribution.

**Figure 7.**
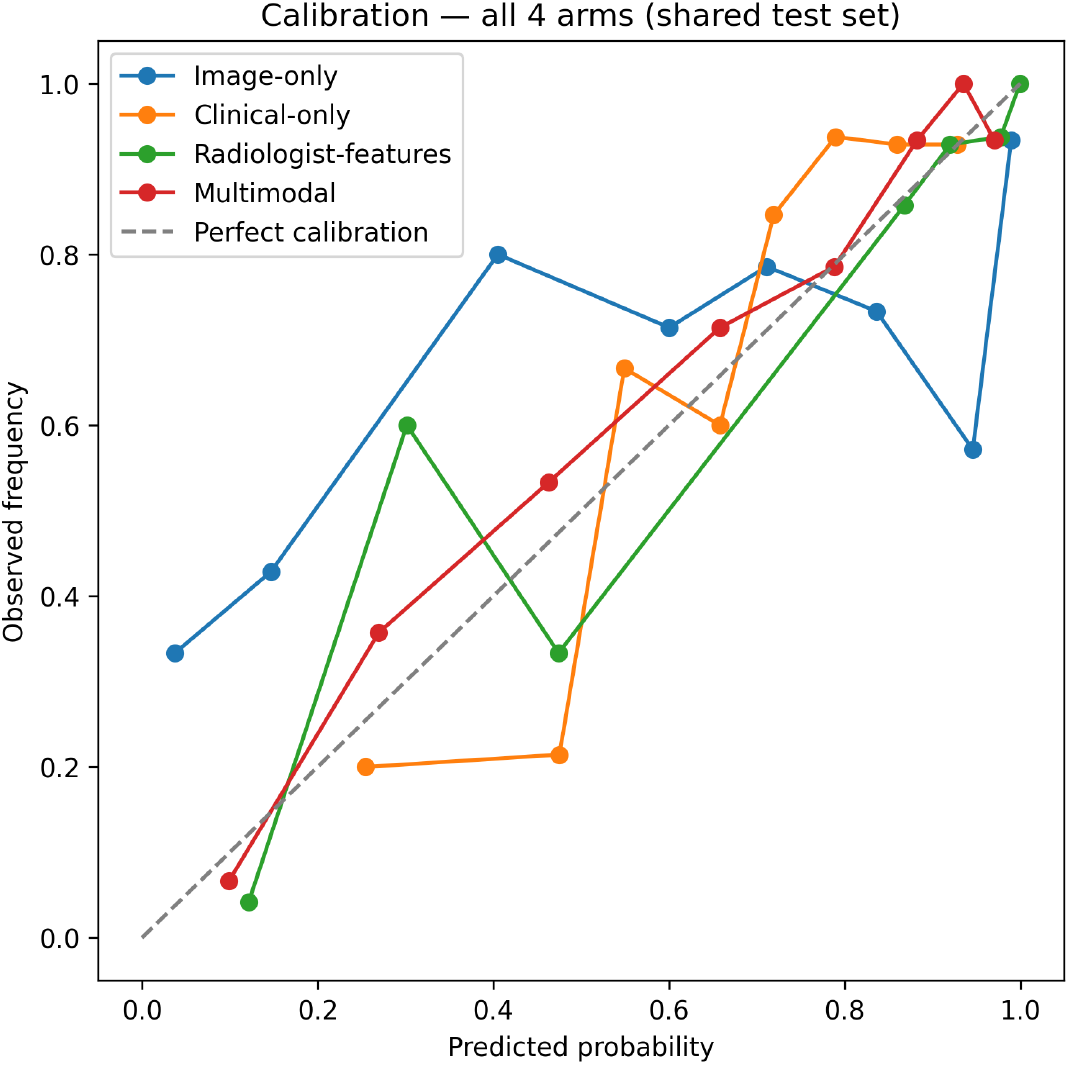
Calibration curves for all four modeling arms on the primary held-out test set.

**Figure 8.**
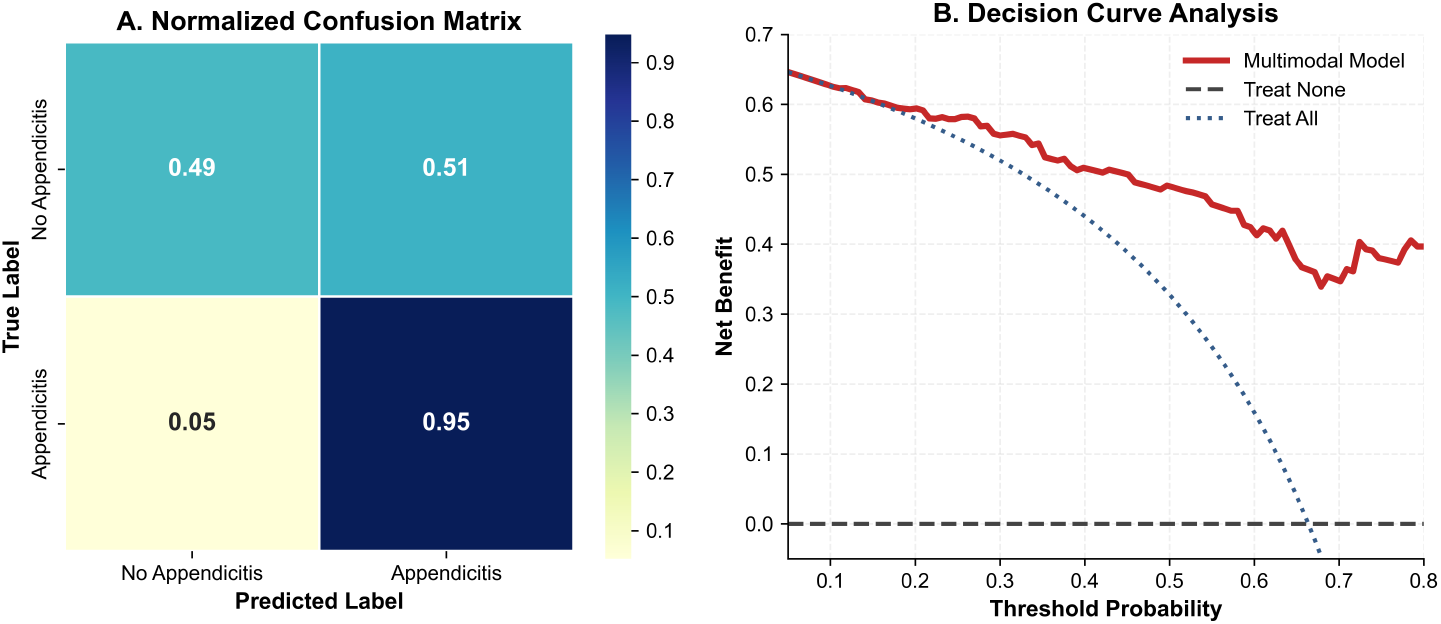
Decision curve analysis for the multimodal model against treat-all/treat-none reference strategies, and a normalized confusion matrix for the multimoal model at its selected operating threshold. The threshold was selected by maximizing Youden’s J statistic on an 80/20 validation split carved out of the 462-patient training set only, then held fixed and applied unchanged to the untouched 116-patient test set; the test set was not used in threshold selectiosn.

## 4 Discussion

### 4.1 Comparison with Prior Work on This Dataset

Table 14 places this study against the three prior analyses discussed in the Introduction. Marcinkevičs et al. [5] built the only other fully automated image pipeline on this dataset, reaching AUROC 0.80 - below our multimodal model’s 0.894, though the two are not directly paired, since the studies used different splits and target formulations. Kendall et al. [6] and Navaei et al. [7] both report near-ceiling performance without processing ultrasound pixel data; the structured fields each calls an “ultrasound” or “imaging” feature include appendix diameter, which we show is partly circular with the outcome for a majority of conservatively-managed patients. We have not seen either study’s code or per-patient predictions and cannot say their specific numbers are inflated by this mechanism; we can say the feature category carries a documented circularity risk on this dataset that neither paper addresses. More broadly, this pattern is consistent with recommendations from systematic reviews of clinical machine-learning studies, which identify insufficiently scrutinized predictor-outcome relationships as a recurring, preventable source of overoptimistic reported performance across the field, not a peculiarity of this dataset [8].

**Table 14.**
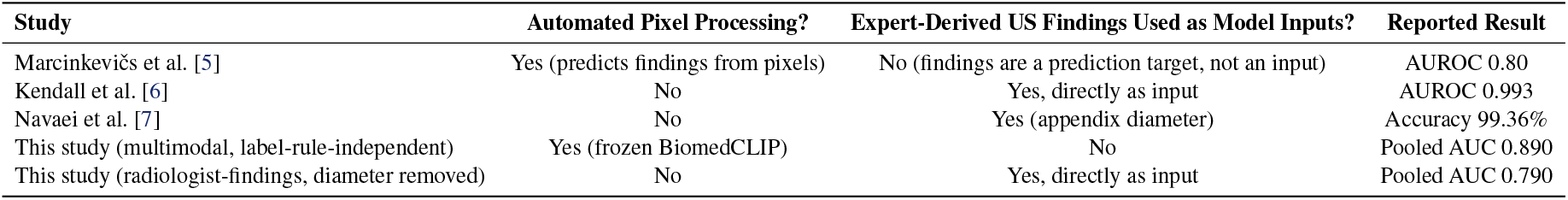
This study in relation to prior work on the Regensburg Pediatric Appendicitis Dataset.

| Study | Automated Pixel Processing? | Expert-Derived US Findings Used as Model Inputs? | Reported Result |
| --- | --- | --- | --- |
| Marcinkevičs et al. [5] | Yes (predicts findings from pixels) | No (findings are a prediction target, not an input) | AUROC 0.80 |
| Kendall et al. [6] | No | Yes, directly as input | AUROC 0.993 |
| Navaei et al. [7] | No | Yes (appendix diameter) | Accuracy 99.36% |
| This study (multimodal, label-rule-independent) | Yes (frozen BiomedCLIP) | No | Pooled AUC 0.890 |
| This study (radiologist-findings, diameter removed) | No | Yes, directly as input | Pooled AUC 0.790 |

### 4.2 Clinical Implications

An automated model using only ultrasound images and routine clinical/laboratory data shows higher discrimination than models built from either input alone, and, once known label-construction dependencies are removed from both sides, also shows significantly higher discrimination than a model built from radiologist-recorded findings. These findings support further evaluation of such a model as a potential decision-support or triage adjunct in settings without immediate access to an experienced pediatric ultrasonographer; they do not establish readiness for clinical deployment. This is a single-center, retrospective, non-externally-validated study with no prospective testing and no comparison against a live radiologist. We report performance by sex and age tertile (Section 3.10) with no marked subgroup deficit observed, but we lack reliable imaging-equipment metadata for a device-level subgroup analysis. All of this would need to be extended before any deployment claim, consistent with what TRIPOD+AI [13] recommends reporting.

### 4.3 Limitations

Several limitations should be stated plainly. The dataset is single-center and retrospective, and the image-linked cohort is capped at 578 patients by what the public release contains; no larger public image archive was identified within the dataset releases we evaluated, though 181 patients with full clinical data have no released image. Every analysis in this study, including the expanded-cohort check, draws on the same institution and the same public dataset family; the study therefore evaluates methodological behavior on a public benchmark rather than demonstrating transportability to independent clinical populations, and no claim of external generalizability should be drawn from it. Our label-construction circularity analysis isolates the contribution of specific predictors but cannot cleanly separate genuine diagnostic signal from labeling-rule dependence within them; the data do not permit separation of the 0.144 AUC decrease observed after removing appendix diameter (Section 3.4) into a circularity-attributable component and genuine diagnostic signal. The image-annotation sensitivity check (Section 3.7), even in its patient-matched form, shows that removing marked images significantly changes model performance, but cannot distinguish whether this is because the models use the displayed numeric measurement itself or because marked and unmarked images differ in framing or quality for other reasons; a proper resolution would require pixel-level masking or inpainting of on-screen measurements while holding the underlying image constant, which we did not implement. Either way, this means the image-only and multimodal models’ reported performance may partly depend on an image-acquisition artifact of this specific dataset rather than transferring unchanged to ultrasound images without such markings. Because of this, our central doubly-corrected comparison (Section 3.6) should not be read as controlling for every known pathway by which expert-derived information could reach the multimodal model: an exploratory check that additionally restricted to annotation-free images (Section 3.8) found a smaller, non-significant difference at the resulting reduced sample size, and we cannot determine from the present data whether this reflects reduced power alone or a genuine additional contribution from the annotation pathway. Our pooled repeated cross-validation reused hyperparameters selected once from the training set across all outer folds rather than re-tuning them in a fully nested design, which may introduce some optimism into the pooled AUC estimates; the magnitude of any such optimism could not be quantified in the present analysis, and a nested re-analysis would be a natural next step. The radiologist-findings model is a retrospective structured-data model, not a real-time radiologist assessment, and should not be read as a claim about live radiologist performance; every comparison in this study, including the doubly-corrected one in Section 3.6, is a model-vs-model comparison and not a direct comparison against human reader performance. We also note that the primary held-out estimates (Section 3.2) and the pooled repeated-cross-validation estimates (Section 3.3) differ somewhat for every arm (for example, multimodal AUC 0.856 vs. 0.894); this reflects that they are estimated from different resampling realizations - a single 116-patient test set versus predictions pooled and averaged across 25 overlapping training/validation splits covering all 578 patients - rather than one estimate being a correction of the other, and both are reported for transparency rather than to be reconciled into a single number. Finally, alternative fusion architectures we tried (backbone and algorithm ensembles; Supplementary Materials) did not improve on the single-backbone multimodal model, suggesting its current pooled AUC may be close to what this dataset and modality combination can support without more data.

## 5 Conclusions

An automated multimodal model built from medical-domain-pretrained ultrasound embeddings and routine clinical/laboratory data showed higher discrimination than both of its unimodal automated counterparts, on a fair, backbone-matched basis, significantly so in pooled repeated cross-validation, on this benchmark. Against a model built from radiologist-interpreted findings, it initially appears to fall short, but we show that both models are affected by outcome-definition dependence through variables entangled in the same labeling rule: the dataset’s rule for labeling non-surgically-managed patients uses appendix diameter, which enters the radiologist-findings model, and two clinical scores, which enter the multimodal model. Correcting for this on both sides - removing appendix diameter from the radiologist-findings model and the two scores from the multimodal model - leaves the multimodal model with significantly higher estimated discrimination under this label-rule-independent, pooled-cross-validation evaluation, though we do not claim this generalizes beyond the evaluated models and validation procedure used here. The practical lesson extends beyond this dataset: wherever structured findings are used both to help construct ground-truth labels for some patients and as input features for a model evaluated against that label, that overlap should be checked and corrected on every arm being compared, not only the one where it is most visible, before a reported accuracy is read as evidence of the model’s diagnostic capability.

## Declarations

### Author Contributions

Conceptualization, methodology, software, formal analysis, data curation, writing-original draft preparation, writing-review and editing, visualization: M.M.I.; supervision: A.Y. The authors have read and agreed to this version of the manuscript.

### Institutional Review Board Statement

Not applicable. This study is a secondary analysis of a publicly available, fully de-identified dataset (the Regensburg Pediatric Appendicitis Dataset [3, 4]). Ethical approval for the original data collection was obtained from the Ethics Committee of the University of Regensburg (approval nos. 18-1063-101, 18-1063_1-101, and 18-1063_2-101), as recorded in the dataset’s public repository entry.

### Informed Consent Statement

Not applicable. This study did not involve direct contact with human subjects; informed consent procedures for the original data collection are described in [5] and the dataset’s repository entry.

### Data Availability Statement

The dataset analyzed in this study is publicly available from Zenodo at https://doi.org/10.5281/zenodo.7669442 (version used) and https://doi.org/10.5281/zenodo.7711412 (latest version, verified identical in image content). Code implementing all models, statistical tests, and sensitivity analyses reported in this study - including the appendix-diameter and Alvarado/PAS circularity analyses, the annotation-contamination check, and the backbone/ensemble ablations - is available at https://github.com/Islam-DS/pediatric-appendicitis-multimodal-ai.

### Use of Artificial Intelligence

During the preparation of this manuscript, M.M.I. used Claude (Anthropic), an AI-based large language model, to assist with data analysis and manuscript drafting. M.M.I. has reviewed, verified, and edited all content, and the authors take full responsibility for the accuracy, integrity, and conclusions of this work.

## Abbreviations

AUC: Area under the receiver operating characteristic curve (used for our own results; AUROC is retained only when quoting a cited study’s own reported terminology)
AP: Average precision
CI: Confidence interval
CRP: C-reactive protein
CNN: Convolutional neural network
DCA: Decision curve analysis
PAS: Pediatric Appendicitis Score
PCA: Principal component analysis
TOST: Two One-Sided Tests
TRIPOD+AI: Transparent Reporting of a multivariable prediction model for Individual Prognosis Or Diagnosis + Artificial Intelligence
WBC: White blood cell

